# Investigating Pathway-Partitioned Polygenic Risk Scores for Schizophrenia: Insights into Clinical Variability in Two Patient Cohorts

**DOI:** 10.64898/2026.04.11.26349671

**Authors:** Jinhan Zhu, Toni A. Boltz, Keith H. Nuechterlein, Robert F. Asarnow, Michael F. Green, Katherine H. Karlsgodt, Diana O. Perkins, Tyrone D. Cannon, Jean M. Addington, Kristen S. Cadenhead, Barbara A. Cornblatt, Matcheri S. Keshavan, Daniel H. Mathalon, William S. Stone, Ming T. Tsuang, Elaine F. Walker, Scott W. Woods, Matthew P. Conomos, Tim B. Bigdeli, Roel A. Ophoff, Carrie E. Bearden, Jennifer K. Forsyth

**Affiliations:** Department of Psychology, University of Washington, Seattle, WA, USA; Stanley Center for Psychiatric Research, Broad Institute of MIT and Harvard, Cambridge, MA, USA; Department of Psychiatry and Biobehavioral Sciences, Semel Institute for Neuroscience and Human Behavior, University of California, Los Angeles, CA, USA; Department of Psychology, University of California, Los Angeles, CA, USA; Department of Psychiatry, University of North Carolina, Chapel Hill, NC, USA; Department of Psychology, Yale University, New Haven, CT, USA; Department of Psychiatry, Hotchkiss Brain Institute, University of Calgary, Calgary, Alberta, Canada; Department of Psychiatry, University of California, San Diego, CA, USA; Department of Psychiatry, Donald and Barbara Zucker School of Medicine at Hofstra/Northwell, Hempstead, NY, USA; Department of Psychiatry, Harvard Medical School at Beth Israel Deaconess Medical Center, Boston, Massachusetts, USA; Department of Psychiatry, University of California, San Francisco, CA, USA; Department of Psychology, Emory University, Atlanta, GA, USA; Department of Psychiatry, Yale University, New Haven, CT, USA; Department of Biostatistics, University of Washington, Seattle, WA, USA; Department of Psychiatry and Behavioral Sciences, Institute for Genomics in Health, State University of New York Downstate Health Sciences University, Brooklyn, NY, USA; Department of Veterans Affairs New York Harbor Healthcare System, Brooklyn, NY, USA; Department of Human Genetics, University of California, Los Angeles, CA, USA; Institute for Public Health Genetics, University of Washington, Seattle, WA, USA; Veterans Affair Greater Los Angeles Health Care System, Los Angeles, CA, USA; Veterans Affairs San Francisco Health Care System, San Francisco, CA, USA

## Abstract

**Background:** Differences in age of psychosis onset (AOO) in schizophrenia (SCZ) are associated with different illness trajectories. Determining whether AOO differences can be explained by genome-wide or pathway-partitioned polygenic risk for SCZ (SCZ-PRS) may elucidate mechanisms underlying clinical variability. This study examined relationships between AOO, genome-wide SCZ-PRS, and pathway-partitioned SCZ-PRS in a harmonized, multi-ancestry North American dataset (SCZ-NA) and in UK Biobank (SCZ-UKBB).

**Methods:** For each cohort, we computed one genome-wide SCZ-PRS and 18 mutually-exclusive pathway-based PRS derived from previous published and validated neurodevelopmental gene-sets. We evaluated 13 SNP-to-gene mapping strategies, including comparing non-coding SNP-to-gene mappings informed by functional annotations versus distance-based windows. SCZ case-control prediction and AOO associations were tested using logistic and linear mixed models, respectively, controlling for sex, ancestry principal components, and genetic relatedness.

**Results:** Genome-wide SCZ-PRS robustly predicted SCZ case-control status in both cohorts but not AOO. In contrast, pathway-based analyses identified AOO associations for a fetal angiogenesis and a postnatal synaptic signaling and plasticity gene-set across both cohorts (*p* < .05), alongside nominal cohort-specific associations in other gene-sets. Associations depended on SNP-to-gene mapping definitions; experimentally informed strategies, particularly those incorporating brain expression Quantitative Trait Locus (eQTL) annotations performed best.

**Conclusion:** Findings suggest that neurovascular and postnatal synaptic signaling and refinement mechanisms contribute to AOO variation in SCZ, and that pathway-informed PRS, especially with brain-specific non-coding SNP-to-gene mappings, can help identify mechanisms contributing to variability in AOO. Replication in larger, prospectively phenotyped cohorts with harmonized AOO definitions will further clarify genetic mechanisms underlying clinical variability in SCZ.

## Introduction

Schizophrenia (SCZ) affects approximately 1% of the global population^[1]^ and is highly heritable (60%-80%)^[2]^. Recent genome-wide association studies (GWAS) have identified hundreds of genetic loci associated with SCZ^[3]^. However, individuals with SCZ carry different risk alleles and also exhibit heterogeneity in clinical characteristics. For example, while psychosis onset commonly occurs in late adolescence to early adulthood^[1]^, it can also manifest as early as childhood^[4]^ or in later adulthood^[5]^. Early-onset psychosis is associated with poorer long-term outcomes, including higher rates of disability and a more chronic illness trajectory^[6–9]^. Understanding whether differences in age of psychosis onset (AOO) reflect distinct genetic profiles has critical implications for risk stratification and the timing of preventive interventions.

Genome-wide polygenic risk scores (PRSs) derived from large-scale GWAS have been shown to explain up to 20% of the variance in SCZ in independent European samples^[10]^. However, PRS often fall short in explaining phenotypic variability^[11]^. For instance, associations between SCZ-PRS and AOO have been inconsistent across studies^[12–15]^. Similarly, findings for SCZ-PRS and cognitive measures are equivocal^[16–22]^. These limitations underscore the need to explore alternate approaches for understanding relationships between genetic and phenotypic variability.

The utility of PRS for explaining clinical variability may be improved by focusing on alleles within specific biological pathways relevant to the disease of interest. Initial pathway-based SCZ-PRS studies suggest that neurotransmitter-specific pathways may relate to cognitive and treatment-related phenotypes in SCZ. Specifically, Pistis et al. (2022) found that an oxidative-stress pathway SCZ-PRS was associated with early psychosis onset, while Warren et al. (2024) found that neurotransmitter pathway-specific SCZ-PRS was associated with deficits in distinct cognitive and functional domains. However, these studies investigated literature-curated, hypothesis-driven gene-sets, which may miss novel mechanisms. They also did not capture neurodevelopmental processes which are central to major etiologic models of SCZ^[25]^. In addition, both studies include overlapping genes across pathways which can create ambiguity, making it difficult to attribute polygenic risk to a specific pathway. In contrast, mutually exclusive co-expression gene sets can be derived in a data-driven way by applying weighted gene co-expression network analysis (WGCNA) to genome-wide transcriptomic data. Prior work has applied WGCNA to subsets of BrainSpan data, which is a large public transcriptomic dataset across the lifespan, to identify genes involved in distinct neurodevelopmental processes^[26,27]^. Using BrainSpan samples from prenatal development through early adulthood, we previously applied WGCNA and identified 18 non-overlapping modules capturing key neurodevelopmental processes^[28]^, such as cell proliferation, neuronal differentiation, myelination, or synaptic signaling and refinement. By focusing on this set of mutually-exclusive neurodevelopmental gene-sets, previously found to have utility for understanding convergent mechanisms of genetic risk for SCZ at the clinical population level, we aimed to test whether polygenic loading across different neurodevelopmental processes relates to phenotypic heterogeneity in SCZ.

Beyond choosing which gene-sets to use for pathway-PRS investigation, an additional critical challenge is determining how best to map non-coding SNPs to genes within these pathways. Most GWAS-associated SNPs are located in non-protein coding regions^[29]^, which often include regulatory elements that influence gene expression across tissues and developmental stages. Thus, optimizing the mapping of non-coding SNPs to genes is essential to optimize pathway-PRS and better understand the genetic mechanisms underlying diseases. One common strategy maps SNPs to genes using fixed windows around gene boundaries^[23,30,31]^. However, enhancer-gene relationships are frequently non-linear and rely on the 3D architecture of the genome for contact, such that many regulatory SNPs do not regulate the nearest gene^[32]^. Relying on nearest-gene, distance-based mapping can therefore attribute regulatory effects to the wrong gene, obscuring molecular mechanisms that underlie disease risk. Addressing these complexities is critical for maximizing the biological accuracy and power of pathway-PRSs for studying SCZ and other complex diseases.

This study therefore aims to determine whether differences in AOO across two SCZ spectrum cohorts can be explained by: (1) genome-wide polygenic risk for SCZ or (2) levels of polygenic risk partitioned within the above-described Brainspan neurodevelopmental pathways^[28]^. In particular, this study will explore the effects of varying definitions for mapping non-coding SNPs to genes when analyzing their contribution within distinct neurodevelopmental pathways to AOO in two SCZ spectrum cohorts. We hypothesized that while genome-wide SCZ-PRS would robustly predict case status, variation in AOO would be better explained by polygenic risk concentrated within specific neurodevelopmental pathways, particularly when SNP-to-gene mapping incorporates brain-relevant functional annotations.

## Materials and methods

### Participants and Clinical Outcome

Analyses were conducted and compared between a cohort of SCZ spectrum patients whose data were harmonized from psychosis-focused research labs across North American (SCZ-NA) and the UK Biobank cohort (SCZ-UKBB).

SCZ-NA included 615 individuals with SCZ and 583 healthy controls, harmonized from multiple studies conducted at UCLA^[33–35]^, nine sites of the North American Prodrome Longitudinal Study (NAPLS), and Zucker-Hillside Hospital in New York^[36,37]^. SCZ-UKBB comprised ∼2700 UK Biobank participants with a SCZ spectrum disorder^[38]^. 20,000 individuals without non-mood psychotic disorders were randomly sampled as controls for SCZ-PRS validation while ensuring computational tractability for the genetic relatedness estimation. In SCZ-NA, SCZ diagnosis and AOO was determined using structured clinical interviews. In primary analysis, SCZ case status included schizophrenia(n=336), schizophreniform disorder(n=92), or schizoaffective disorder(n=49)^[39]^. In SCZ-UKBB, diagnoses were ascertained from self-report and linked health records; SCZ case status included schizophrenia or schizoaffective disorder; no UKBB subjects had diagnoses of schizophreniform disorder. AOO was defined from self-report data. See Supplementary Methods for details.

### Genotyping and imputation

SCZ-NA samples were genotyped in two batches on the Illumina Global Screening Array, and underwent standard pre- and post-imputation quality control using RICOPILI^[40]^. Imputation was performed using the Trans-Omics for Precision Medicine (TOPMed) reference panel^[41–43]^. SCZ-UKBB analyses were based on imputed genotype data that passed UK Biobank quality control procedures^[44]^. In both cohorts, multi-allelic sites and variants in the extended major histocompatibility complex region were excluded except the SNP showing strongest association with SCZ^[3,10]^. See Supplementary Methods for details.

### Genome-wide and pathway-specific polygenic risk scores

For each cohort, we computed one genome-wide SCZ-PRS. For pathway-based SCZ-PRS, we used the 18 mutually-exclusive neurodevelopmental gene-sets described above^[28]^. Within each gene set, we applied 13 distinct SNP-to-gene mapping definitions that differed along two dimensions: how variants within gene boundaries were defined and how non-coding variants outside gene bodies were assigned to genes. For variants between a gene’s transcription start and end sites, we considered two genic filters: 1) an Exon + Splice definition restricted to ANNOVAR-annotated^[45]^ exonic and splicing categories, and 2) an Exon + Intron definition including all variants within the transcribed region, including both exonic and intronic variants. Non-coding variants outside gene bodies were then linked to genes using either distance-based window approaches or functional annotation-based mappings (Figure 1). Functional mappings incorporated brain-specific expression and splicing quantitative trait loci (eQTL/sQTLs) data^[46,47]^, enhancer-promoter annotations^[48]^, and/or fetal brain chromatin contacts^[49]^. See Supplementary Methods for detailed mapping definitions.

**Figure 1.**
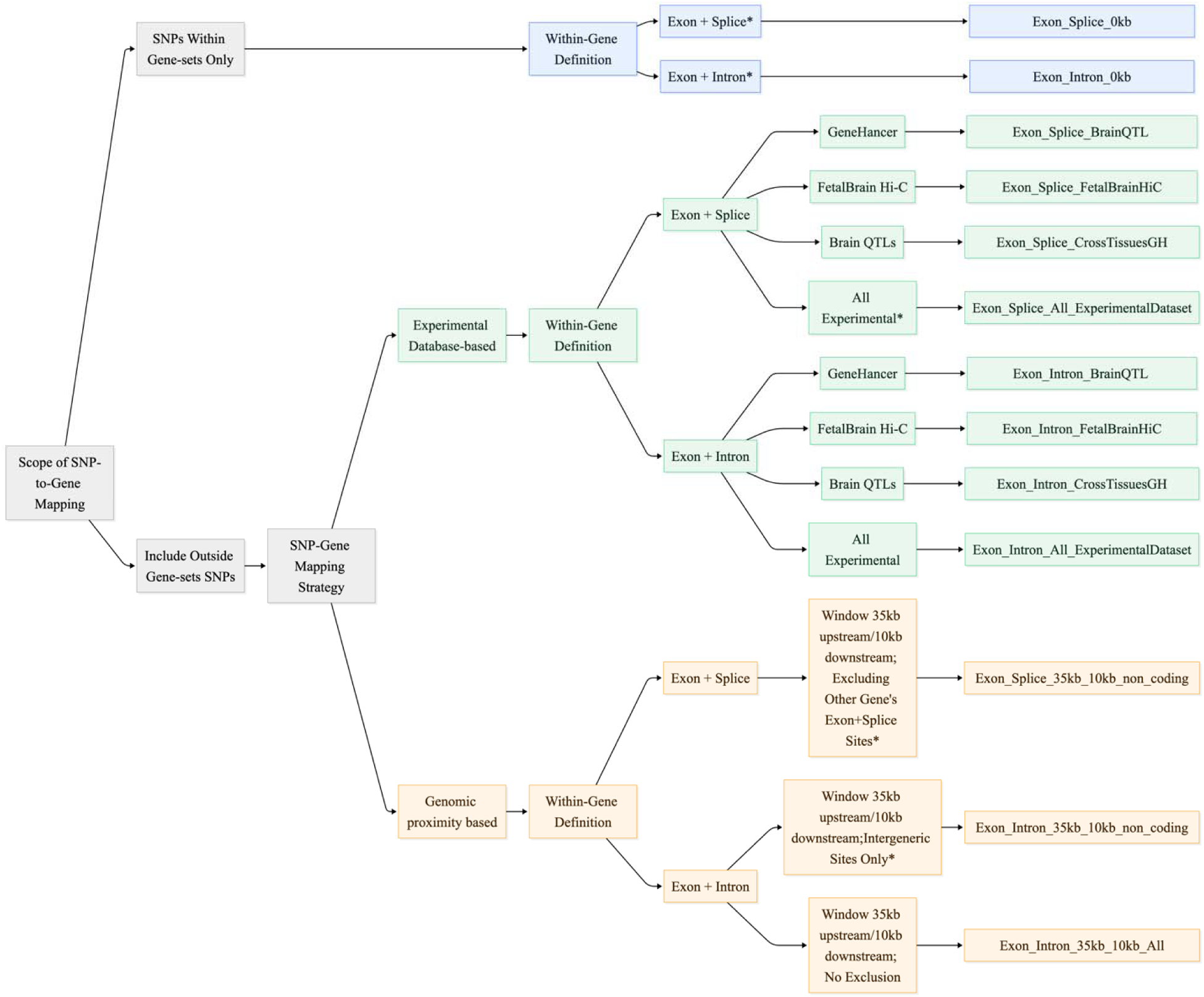
SNP-to-Gene Mapping Workflow. Schematic of the hierarchical framework used to define SNP-gene mappings in pathway-based PRS analyses. SNPs were either restricted to located within the transcription start to end of gene-sets genes only or extended to include SNPs located outside the gene bodies of the target gene-sets. For SNPs located within gene boundaries, inclusion criteria were defined using either “Exon + Splice” or “Exon + Intron” annotations. For strategies extending beyond gene boundaries, SNP-to-gene assignment was based on either experimental datasets or genomic proximity 35-kb upstream and 10-kb downstream with additional genic functional-annotation filtering. The rightmost column lists the abbreviations for the resulting 13 mapping definitions used in our analyses. (1) *Exon + Splice* = variants annotated as “splicing,” “ncRNA_exonic;splicing,” “exonic;splicing,” “ncRNA_exonic,” “ncRNA_splicing,” or “exonic”; (2) *Exon + Intron* = includes all variants located within the gene body; (3) *All Experimental* = union of GeneHancer, Brain QTL, and Fetal Hi-C datasets; (4) *Excluding Other Gene’s Exon + Splice Sites* = for SNPs outside 35-kb/10-kb window size, removes those located within other genes’ exonic or splicing regions; (5) *Intergeneric Sites Only* = for SNPs outside 35-kb/10-kb window size, only keep SNPs annotated as “intergenic,” “upstream,” “downstream,” or “upstream;downstream.”

### Polygenic risk scores calculation and ancestry adjustment

Given the SCZ-NA cohort’s multi-ancestry composition, we employed PRS-CSx^[50]^, which improves cross-population PRS transfer by leveraging ancestry-specific LD patterns. We used predominantly European-ancestry summary statistics from the PGC3 SCZ GWAS^[3]^, alongside smaller SCZ GWAS of Admixed American and African ancestry individuals^[51]^. The inverse-variance-weighted meta-analysis of population-specific posterior SNP estimates were used to construct SCZ-PRS. Because SCZ-UKBB is composed predominantly of individuals of European descent, we derived SCZ-PRS using PGC3 SCZ GWAS^[3]^ and applied PRS-CS^[52]^, a Bayesian approach that accounts for LD structure and is well suited to ancestry-matched European target cohorts^[50]^. In both cohorts, posterior SNP weights from PRS-CS/PRS-CSx were allele-aligned to the target genotypes and per-individual scores were computed in PLINK v2.0. To improve comparability across ancestry backgrounds, all PRSs were post hoc ancestry-adjusted using a continuous PCA-based framework anchored to 1000 Genomes reference data^[53]^, which models ancestry-related differences in both the mean and variance of PRS distributions^[54]^. See Supplementary Methods for details.

### Statistical analysis

Logistic regression models were used to validate the association between genome-wide PRS with SCZ case-control status. Specifically, SCZ case-control status was regressed on the genome-wide PRS, with sex, age at assessment, top 10 ancestry PCs, and the genetic relatedness matrix as covariates. Linear mixed models were then used to test associations between genome-wide SCZ-PRS and pathway-specific SCZ-PRS with AOO, with the same covariates except age at assessment (i.e., which was partially confounded with AOO). Analyses were conducted using the GENESIS package in R^[55]^, which allows robust modeling of genetic data while accounting for genetic relatedness. For each pathway PRS, we computed a competitive, permutation-based p-value to quantify enrichment beyond what would be expected for a random SNP set of comparable size. For each gene set, we generated 10,000 null PRSs by randomly sampling the same number of non-zero weighted SNPs as in the pathway PRS from the genome-wide SNP pool, output from PRS-CS or PRS-CSx. For each null PRS, we fit the same association model and recorded the association statistic. Following the PRSet method^[30]^, the competitive p-value was defined as the proportion of null PRSs whose association strength exceeded that of the observed pathway PRS. This approach accounts for differences in SNP count across gene-set definitions.

## Results

### Associations of Genome-wide SCZ-PRS with diagnostic status

Sample demographics and clinical characteristics for both cohorts are presented in Table 1. In the SCZ-NA cohort, PRS-SCZ explained 6.8% of the variance in case-control status (p = 3.309E-16), with individuals in higher PRS quintiles showing progressively elevated odds of case status relative to the lowest quintile (Figure 2A). Similarly, in the SCZ-UKBB cohort, PRS-SCZ explained 3.5% of the variance in SCZ case-control status (p = 2.102E-169); individuals in the top quintile exhibited nearly a 5-fold higher odds of SCZ status compared with those in the bottom quintile (Figure 2B).

**Figure 2.**
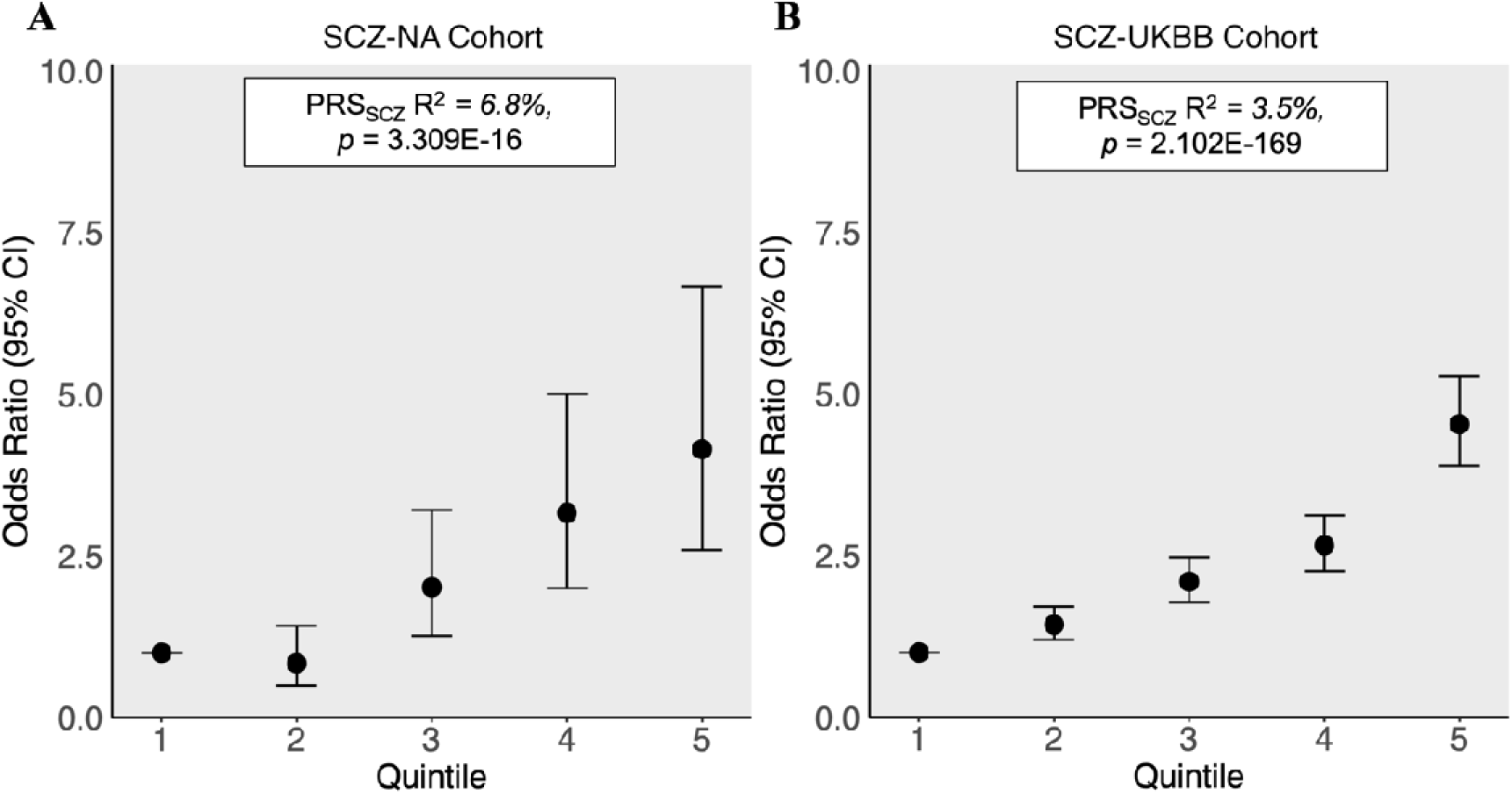
Genome-wide SCZ-PRS and schizophrenia case status across quintiles. Odds ratios (OR) and 95% CIs for SCZ case status by genome-wide SCZ-PRS quintile (Q1-Q5), with Q1 as reference in the (A) SCZ-NA; and (B) SCZ-UKBB cohorts. Insets show variance explained (R²) and P values for the genome-wide PRS in each cohort. A monotonic increase in OR across quintiles is evident in both cohorts, demonstrating robust SCZ case-control discrimination by the genome-wide SCZ-PRS.

**Table 1.** Demographic and Clinical Characteristics of the SCZ-NA and SCZ-UKBB Cohorts.

|  | SCZ-NA |  | SCZ-UKBB |  |
| --- | --- | --- | --- | --- |
| SCZ Status | SCZ Spectrum | Controls | SCZ Spectrum | Controls |
| Subjects N (% Female) | 609 (30.2%) | 581 (50.4%) | 2668 (48.4%) | 19722 (54.1%) |
| Mean Age ± SD | 23.85 ± 10.3 | 24.59 ± 12.3 | 55.82 ± 8.3 | 56.55 ± 8.11 |
| N SCZ Diagnosis | 477 | NA | 1390 | NA |
| Schizophrenia | 336 |  | 1353 |  |
| Schizophreniform | 92 |  | 0 |  |
| Schizoaffective | 49 |  | 37 |  |
| N SCZ with AOO Information | 471 | NA | 625 | NA |
| Mean AOO ± SD | 20.02 ± 6.0 | NA | 32.0 ± 12.9 | NA |
| Race (%) of Participants with AOO Data |  |  |  |  |
| African American/Black | 119 (25.3%) |  | 7 (1.1%) |  |
| Asian | 48 (10.2%) |  | 23 (3.7%) |  |
| Caucasian/White | 163 (34.6%) |  | 593 (94.9%) |  |
| Mixed/Other | 141 (29.9%) |  | 2 (0.4%) |  |

*Note.* AOO = age of psychosis onset.

### Associations of Genome-wide SCZ-PRS with Age of Psychosis Onset

We next examined the association between genome-wide SCZ-PRS and AOO across cohorts. As shown in Figure 3, genome-wide PRS was not significantly associated with AOO in the SCZ-NA cohort (b=0.44, p=.081) or the SCZ-UKBB cohort (b=−0.26, p=.595). Although non-significant, the direction of effect differed across samples. Individuals with higher SCZ-PRS in the SCZ-NA cohort, which had an earlier than average AOO overall^[39]^, tended to have a later AOO. Conversely, in the SCZ-UKBB cohort, which had a later than average AOO overall^[56]^, individuals with higher SCZ-PRS tended to have an earlier AOO.

**Figure 3.**
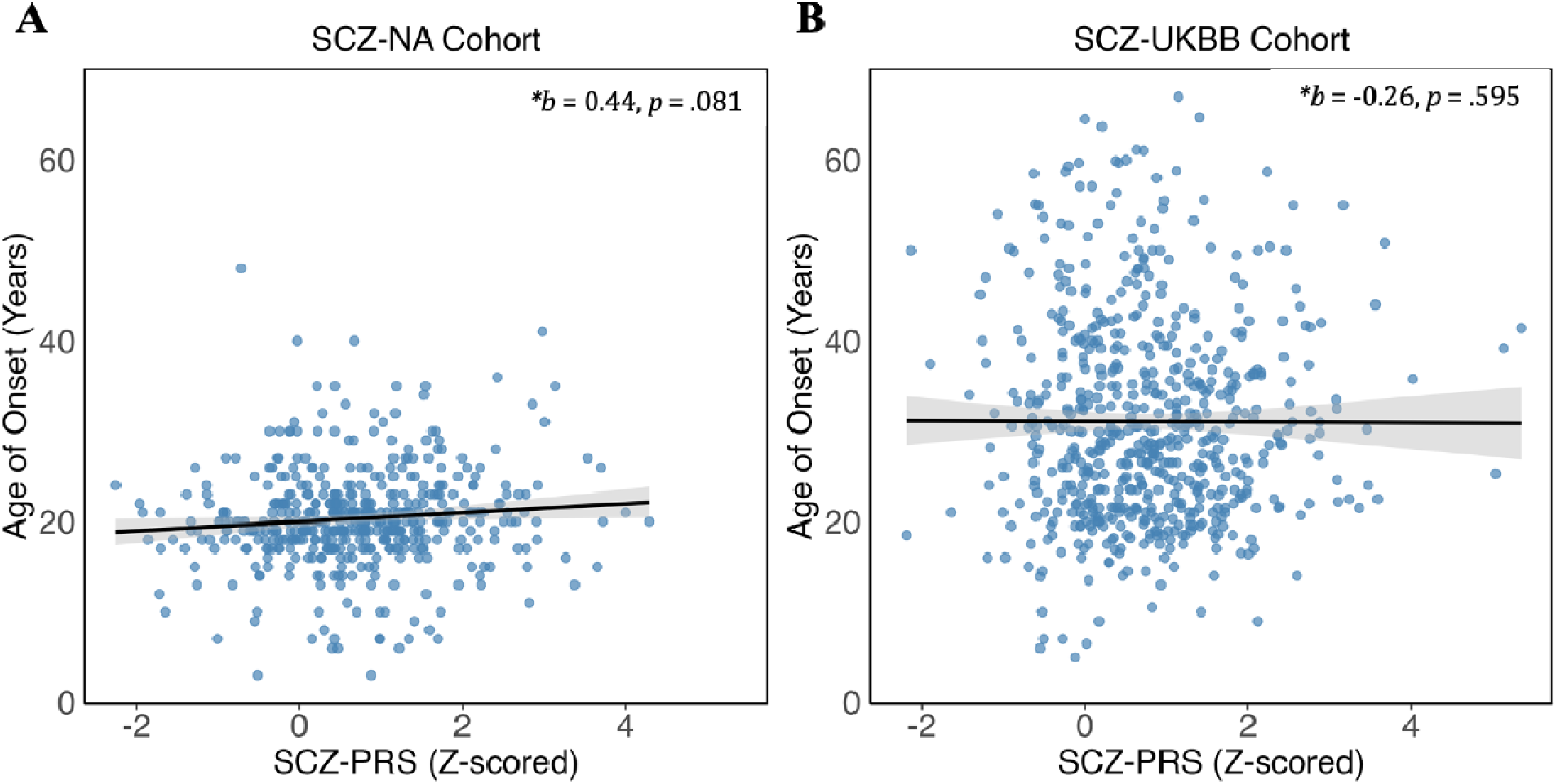
Genome-wide SCZ-PRS and AOO. Associations between z-scored genome-wide SCZ-PRS and AOO (years) in the (A) SCZ-NA and (B) SCZ-UKBB cohorts. Scatterplots report the regression coefficient (b) and p-value for the SCZ-PRS term in each cohort. No significant association was observed in either cohort, and effect directions differed across samples.

### Pathway SCZ-PRS SNPs Assignment

To evaluate whether neurodevelopmental pathway-partitioned polygenic profiles provide insight into AOO variability, we constructed pathway-specific PRS and tested their associations with AOO using the SNP-to-gene mapping definitions summarized in Figure 1.

A detailed breakdown of SNP counts for each gene-set under each SNP-to-gene mapping definition is provided in Supplementary Table 1. Gene-set-specific mean SNP counts across SNP-to-gene mapping definitions are reported in Supplementary Table 2, and SNP-to-gene mapping definition-level mean SNP counts across gene-sets are summarized in Supplementary Table 3. The most restrictive SNP-to-gene definition was Exon_Splice_0kb, yielding a mean SNP count of 920 and 1122 across gene-sets in SCZ-NA and SCZ-UKBB, respectively. In contrast, the broadest annotations include intronic variants plus either all regulatory datasets (i.e., “Exon_Intron_All_ExperimentalDataset”; mean SNP numbers across gene-sets: SCZ-NA=33868; SCZ-UKBB=49270) or the 35-kb upstream/10-kb downstream window without functional-annotation filtering (i.e., “Exon_Intron_35kb_10kb_All”; mean SNP numbers across gene-sets: SCZ-NA=28291;SCZ-UKBB=40709).

### Associations of Pathway-based SCZ-PRS with Age of Psychosis Onset

Results of association analyses for AOO are presented in Figure 4 and Supplementary Table 4. For each gene-set within each cohort, we identified the “best-performing” SNP-to-gene mapping definition as the one explaining the most variance. Among the 36 best-performing definitions across cohorts, 26 applied the “Exon+Splice” genic filter, and 20 incorporated experimentally-derived regulatory information. Variants of the commonly used 35-kb upstream/10-kb downstream window appeared less often among the top-performing definitions, and all distance-based best performers excluded SNPs located within the genic regions of genes not included in the focal gene set. Among gene-sets showing nominal signal associations, 9 of 10 applied the “Exon+Splice” filter, and among the 7 that included non-coding SNPs, 6 incorporated experimentally derived regulatory information versus 1 used distance-based information. Overall, SNP-to-gene mapping definitions providing the highest variance explained generally prioritized genic functional filtered SNP assignment and often incorporated more precise experimentally derived regulatory annotations, whereas distance-based window approaches less frequently provided the highest variance explained, particularly among gene-sets showing nominal signal associations.

**Figure 4.**
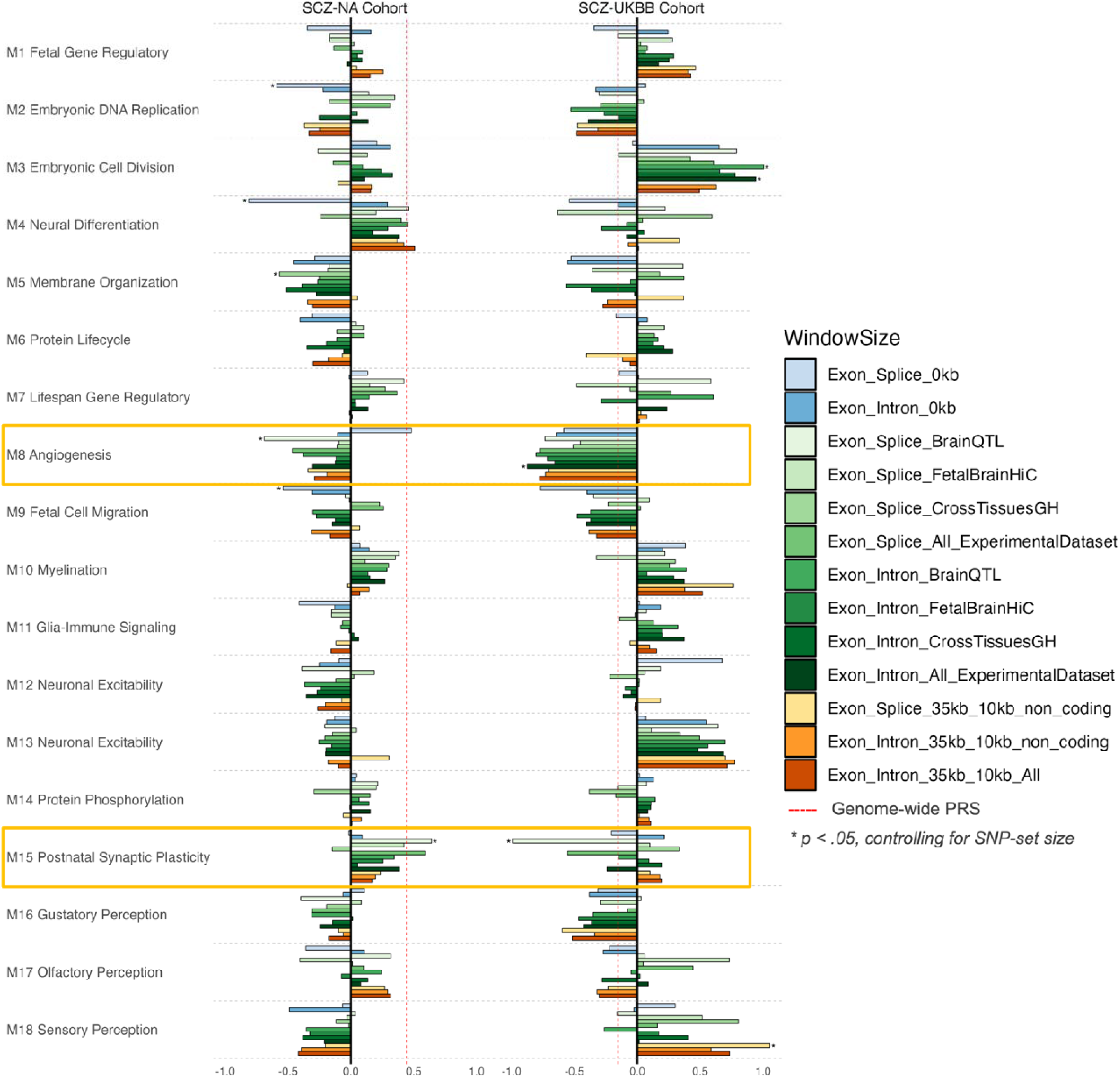
Pathway-based SCZ-PRS associations with AOO across SNP-gene mapping strategies. Bars show the regression coefficient (b) from mixed linear models of AOO on pathway-specific PRS, for (left) SCZ-NA and (right) SCZ-UKBB cohorts. Negative *b* indicates that higher pathway PRS is associated with earlier AOO while positive *b* indicates association with later AOO. Colors denote 13 SNP-gene mapping definitions (right legend), grouped as within-gene SNPs only (blue) and non-coding SNP-to-gene mappings based on experimental annotations (green) or 35-kb upstream / 10-kb downstream proximity (orange). The red dashed line marks the corresponding genome-wide SCZ-PRS effect for reference. * *p* < .05 after controlling for SNP-set size. Gene-sets with replicated signals (M8 and M15) are highlighted in yellow boxes.

Nominal pathway-level SCZ-PRS associations were observed after accounting for pathway SNP counts (Table 2). Although no associations survived multiple-comparisons correction across pathways and mapping definitions, two neurodevelopmental gene-sets were notable in demonstrating associations with AOO across both cohorts. The M15 gene-set involved in postnatal synaptic signaling and plasticity was associated with AOO specifically under the “Exon_Splice_BrainQTL” SNP-to-gene mapping definition for both cohorts (SCZ-NA: maximum R²=1.3%, competitive-p=.0244; SCZ-UKBB: maximum R²=0.7%, competitive-p=.0247). The direction of effect differed between cohorts: higher postnatal synaptic signaling SCZ-PRS was associated with later onset in the SCZ-NA cohort, but with earlier AOO in the SCZ-UKBB cohort. The M8 gene-set involved in fetal angiogenesis also showed associations with AOO in both cohorts (SCZ-NA, maximum R²=1.3%, p=0.0175 under the “Exon_Splice_BrainQTL” SNP-to-gene mapping definition; SCZ-UKBB, maximum R²=0.7%, p=0.0318 under the “Exon_Intron_All_ExperimentalDataset” SNP-to-gene mapping definition). In both cohorts, higher loading of risk variants in the fetal angiogenesis SCZ-PRS was associated with earlier psychosis onset.

**Table 2.** Nominal pathway-level SCZ-PRS associations with AOO,corrected by gene-set size.

| Neurodevelopmental.Gene-Sets | Cohorts | SNP-to-Gene Definition | Effect Size | Competitive_P |
| --- | --- | --- | --- | --- |
| M2 Embryonic DNA Replication | SCZ-NA | Exon_Splice_0kb | -0.5911 | 0.0312 |
| M4 Neural Differentiation | SCZ-NA | Exon_Splice_0kb | -0.8127 | 0.0011 |
| M5 Membrane Organization | SCZ-NA | Exon_Splice_CrossTissuesGH | -0.5683 | 0.0309 |
| M8 Angiogenesis | SCZ-NA | Exon_Splice_BrainQTL | -0.688 | 0.0175 |
| M9 Fetal Cell Migration | SCZ-NA | Exon_Splice_0kb | -0.5406 | 0.037 |
| M15 Postnatal Synaptic Plasticity | SCZ-NA | Exon_Splice_BrainQTL | 0.6407 | 0.0244 |
| M3 Embryonic Cell Division | SCZ-UKBB | Exon_Splice_BrainQTL | 1.0044 | 0.0228 |
| M8 Angiogenesis | SCZ-UKBB | Exon_Intron_All_ExperimentalDataset | -0.8749 | 0.0318 |
| M15 Postnatal Synaptic Plasticity | SCZ-UKBB | Exon_Splice_BrainQTL | -0.9914 | 0.0247 |
| M18 Sensory Perception | SCZ-UKBB | Exon_Splice_35kb_10kb_non_coding | 1.0543 | 0.013 |

*Note.*
AOO=age of psychosis onset.
Competitive\_P: The P-value correcting for gene-set size and SNP density by comparing the association of
the target gene-set PRS against a null distribution generated from 10,000 random gene-sets containing an equivalent number of SNPs.

In addition to cross-cohort associations, several nominal associations emerged that were specific to individual cohorts. Within the SCZ-NA cohort, pathway PRS for an embryonic DNA Replication gene-set (M2), a neuronal differentiation gene-set (M4), a membrane organization gene-set (M5), and a fetal cell migration gene-set (M9) were each associated with earlier AOO (b range=−0.8127 to −0.5406, all competitive-p < 0.05; Figure 4), whereas in the SCZ-UKBB cohort, pathway PRS for an embryonic cell division gene-set (M3) and a sensory perception gene-set (M18) were associated with later AOO (b range=0.9430 to 1.0543, all competitive-p < 0.05; Figure 4), depending on SNP-to-gene mapping definition. Across the six nominally significant associations, five were observed when using an “Exon+Splice” functional filter, suggesting that more functionally constrained and empirically informed SNP-to-gene mappings may reveal stronger pathway-level genetic risk loading relationships with AOO.

## Discussion

In this study, we move beyond conventional genome-wide polygenic scoring to test whether biologically informed neurodevelopmental pathway-partitioned SCZ-PRS captures variation AOO in SCZ. Across two independent cohorts, genome-wide SCZ-PRS robustly predicted diagnostic status but showed no association with AOO, suggesting limited sensitivity of aggregate polygenic burden for this clinical feature. In contrast, pathway-level analyses identified two neurodevelopmental gene-sets associated with AOO across cohorts: a postnatal synaptic signaling and plasticity gene-set (M15) and a fetal angiogenesis gene-set (M8). In addition, several nominal associations emerged that were cohort-specific. Importantly, SNP-to-gene mapping strategy influenced associations between pathway-specific SCZ-PRS and AOO. For the M8 and M15 gene-sets that showed associations across cohorts, associations arose from experimentally-informed mappings, particularly those incorporating brain eQTL. These findings suggest that while genome-wide SCZ-PRS does not robustly relate to variation in AOO, biologically-informed polygenic profiles may reveal mechanistic insights.

### Genome-wide SCZ-PRS and clinical phenotypes

Consistent with prior work demonstrating robust associations between genome-wide SCZ-PRS and SCZ case-control status^[3,57]^, our validation analyses showed that the genome-wide PRS strongly predicted SCZ diagnostic status. The association between SCZ-PRS and case-control status in SCZ-NA (OR=1.70) was comparable to prior European-ancestry reports, including Crouse et al. (2021) for psychotic disorder (OR=1.68) and strict SCZ case-control analyses from PsycheMERGE consortium (OR=1.55)^[59]^ and the U.S. Veterans Affairs system (OR=1.56)^[60]^. In the SCZ-UKBB cohort, the association was also consistent with previous UK Biobank findings across ancestries^[56]^ (OR=1.69) with our analysis yielding a modestly higher odds ratio (OR=1.89). Variation in effect sizes between our cohorts and previous studies may be attributable to methodological differences, including SCZ case ascertainment, inclusion of more diverse ancestry groups with dimensional ancestry adjustment, and analytical correction for genetic relatedness.

### Genome-wide SCZ-PRS and AOO

In the present study, genome-wide SCZ-PRS was not associated with AOO in either cohort. This is consistent with several prior studies reporting no significant association between SCZ-PRS and AOO^[12–14]^. One prior study of ∼4,740 European-ancestry participants reported a small but significant association between SCZ-PRS and earlier AOO (R²=0.13%)^[15]^. That study employed a broader psychosis-spectrum case definition, including delusional disorder, brief psychotic disorder, and psychotic disorder not otherwise specified. This is relevant because AOO differs across psychotic diagnoses, with SCZ typically emerging in late adolescence or early adulthood, whereas delusional disorder more often has a later onset^[61]^. Prior work also suggests that SCZ polygenic liability varies across psychotic diagnoses and symptom-defined subgroups^[62]^, raising the possibility that this broader case definition may dilute schizophrenia-specific associations with age at onset. By contrast, our study leveraged the largest and most up-to-date SCZ GWAS to construct SCZ-PRS^[3]^, applied a commonly used and clinically grounded definition of AOO, and included cohorts with more diverse ancestry backgrounds, thereby enhancing statistical power and generalizability. Together, our findings add to a growing body of literature suggesting that genome-wide SCZ-PRS has minimal association with AOO among individuals with a narrower set of psychotic diagnoses.

### Pathway-specific SCZ-PRS and AOO

While genome-wide SCZ-PRS was not associated with AOO, pathway-specific SCZ-PRSs, partitioned based on neurodevelopmental gene-sets, suggest novel insights into mechanisms underlying AOO differences. Higher SCZ risk loading within a postnatal synaptic signaling and plasticity gene-set (M15) was nominally associated with AOO in both the SCZ-NA and SCZ-UKBB cohorts, but in opposite, “centralizing” directions. This directional difference likely reflects cohort-specific ascertainment and phenotype composition. SCZ-NA is enriched for individuals with earlier and recent-onset psychosis, including individuals with child- or adolescent-onset psychosis^[39]^, whereas SCZ-UKBB is drawn from a population biobank of adults with a comparatively later AOO distribution. The M15 gene-set was previously found to be strongly enriched for risk variants for SCZ, including both common variants and copy number variants^[28]^. Functionally, M15 is enriched for neuronal markers and genes involved in neurotransmitter release, regulation of neurotransmission, and learning and memory, with several hub genes linked to synaptic development and neuropsychiatric risk. Notably, expression of M15 increases postnatally until early adulthood^[28]^. Within a neurodevelopmental framework of SCZ, disruption of M15 genes may therefore exert its strongest effects during late adolescence and early adulthood, when synaptic refinement and maturation of excitatory-inhibitory balance extend into late-maturing brain regions implicated in SCZ onset^[63]^. These findings suggest that higher polygenic burden in a canonical SCZ signaling pathway may shift AOO toward the typical window of late adolescence and early adulthood, pointing to postnatal synaptic signaling and plasticity as a potential key biological substrate influencing the developmental timing of symptom onset in SCZ.

A fetal angiogenesis gene-set (M8) also showed nominal associations with earlier AOO in both cohorts. M8 is enriched for angiogenic and developmental growth processes and demonstrates prenatal-biased expression, suggesting neurovascular mechanisms in schizophrenia onset. In a recent Dynamic Contrast-Enhanced MRI study of 41 individuals with SCZ, widespread increases in blood-brain barrier (BBB) leakage were detected across regions typically affected in SCZ, providing the first in-vivo evidence of subtle barrier disruption in SCZ^[64]^. BBB dysfunction can amplify neuroimmune cascades by permitting peripheral cytokines and immune cells to enter the brain tissues, activating microglia and astrocytes and further compromise neuronal and synaptic integrity. Although speculative, perturbations in fetal angiogenesis could hinder BBB integrity and neurovascular homeostasis, thereby heightening vulnerability to earlier onset^[65]^. Together, the cross-cohort convergence of M8 and M15 suggests that both neurovascular and synaptic signaling mechanisms may be involved in the developmental timing of psychosis onset.

Several additional PRS-pathway associations with AOO were identified that were cohort-specific. Specifically, gene-sets related to embryonic DNA replication (M2), neuronal differentiation (M4), membrane organization (M5), and fetal cell migration (M9) were associated with earlier onset in the SCZ-NA cohort. M4 and M5 are both enriched for neuronal cell-type markers. These nominal associations suggest that disruption of neuronal development may contribute to earlier clinical emergence. Conversely, in SCZ-UKBB, M3, involved in cell-cycle control of progenitor cells, and M18, a sensory perception and receptor-signaling gene-set, were associated with later AOO in SCZ-UKBB. The precise mechanisms through which altered cell-cycle parameters contribute to later psychosis onset are unclear, but risk alleles in genes involved in sensory perception suggest that differences in neural processing of environmental stimuli could contribute to variation in AOO. Together, these cohort-specific patterns suggest that psychosis onset may reflect the cumulative impact of genetic variation across multiple neurodevelopmental windows while differences across cohorts may also reflect ascertainment heterogeneity and measurement variation. Replication is required in larger samples.

### SNP-to-Gene Mapping Definition

Our findings contribute to the debate over how best to annotate non-coding SNPs to genes to capture true regulatory relationships, an area of active research. Many studies have historically relied on distance-based definitions, in which SNPs are linked to genes using a user-selected window size outside the gene body. However, there is substantial biological ambiguity with this approach^[66]^. To our knowledge, this is the first study to systematically evaluate multiple mapping definitions within one framework. The observation that experimentally informed mappings, particularly those incorporating brain eQTLs, yielded the most consistent signals highlights the potential value of functionally grounded approaches. These results align with a shift in polygenic prediction toward integrating functional information beyond GWAS summary statistics. For example, Zheng et al. (2024) recently introduced a polygenic prediction framework that integrates genome-wide imputed SNPs with functional annotations to hone in on potential causal SNPs relevant for polygenic prediction, yielding promising increases in PRS accuracy in diverse populations^[67,68]^. Recent pathway-based PRS studies have also increasingly moved in this direction, integrating functional annotations and experimentally derived datasets such as brain eQTLs and chromatin interaction maps to refine SNP-gene mapping and enhance biological relevance^[23,31,69,70]^. To our knowledge, only one prior study compared SNP-to-gene assignment strategies in a pathway-partitioned SCZ-PRS framework; this study observed an association between an oxidative stress pathway SCZ-PRS and early psychosis only when mapping was based on eQTLs rather than distance-based definitions^[23]^. Our findings suggest that functional annotation-informed mapping strategies better capture regulatory biology and maximize accuracy of pathway-PRS for patient stratification.

There are several limitations to the current study that should be acknowledged. Partitioning polygenic risk into distinct pathways, while enhancing biological interpretability, may oversimplify crosstalk and synergistic effects among pathways. The modest effect sizes and nominal significance of our associations also underscore the need for replication in larger, well-powered cohorts. The use of two independent datasets, although valuable for validation, also introduced heterogeneity in AOO assessment, genotyping procedures, and the cohorts’ age and ancestry composition, which may affect cross-cohort comparability. A specific limitation of the SCZ-UKBB cohort is that AOO was based on retrospective self-report, raising the possibility of recall bias that could affect estimates of onset timing and the robustness of the findings. Moreover, while the 18 Brainspan neurodevelopmental gene-sets investigated here were biologically validated previously^[28]^, pathway PRS using alternative literature-curated or co-expression-derived gene-sets may reveal other mechanistic insights^[71]^. Future work should expand beyond these gene-sets and leverage larger datasets with harmonized AOO definitions that can accommodate stricter multiple-testing correction to identify the optimal biological parcellation of PRS for patient stratification.

In summary, this study introduces a pathway-partitioned polygenic risk framework for dissecting genetic contributions to heterogeneity in SCZ AOO. By integrating neurodevelopmentally-informed gene-sets with multiple SNP-to-gene mapping strategies, our findings suggest that biologically grounded PRS-partitioning can uncover mechanistic dimensions of risk that are obscured in genome-wide PRS approaches. Our findings also suggest that tissue-specific functional annotations and experimentally derived links, especially brain QTL, tend to outperform distance-based SNP-to-gene mapping for capturing regulatory regions. Replicable cross-cohorts associations in fetal angiogenesis (M8) and postnatal synaptic signaling and plasticity (M15) pathways, in particular, suggest important neurodevelopmental routes to illness onset. These findings suggest that integrating pathway-specific polygenic models can help refine patient stratification and could eventually contribute to more mechanistically informed approaches to early intervention in schizophrenia.

## Acknowledgements

This research has been conducted using the UK Biobank Resource under Application Number 136530. This work was supported by NIMH (K08 MH118577 to JKF; MH101506 to KHK; R01 MH37705, R01 MH37705, R01 MH110544, P50 MH066286, and R01 MH130848 to KHN; U01 MH082004 to DOP; U01 MH081902 to TDC; U01 MH081988 to EFW; NIMH U01 MH081902 , P50 MH066286 , R01MH123641, and Levin Trust to CEB; the Brain & Behavior Research Foundation (NARSAD Young Investigator Award to JKF), the National Center for Advancing Translational Sciences UCLA CTSI Grant UL1TR001881 (Award to CEB & JKF), the UCLA Brain Research Institute (Postdoctoral Award to JKF), and the Shear Family Foundation.

## Data Availability

This study used data from two cohorts: a harmonized, multi-ancestry North American cohort and UK Biobank. Data from the harmonized, multi-ancestry North American cohort used in this study, collected under informed consent procedures consistent with broad data sharing, are available through NIH Data Archive Collection #3226. UK Biobank data are available through the UK Biobank data access procedures.

