## Supplementary Methods for "Investigating Pathway-Partitioned Polygenic Risk Scores for Schizophrenia: Insights into Clinical Variability in Two Patient Cohorts"

**SUPPLEMENTARY INFORMATION**

### **Supplementary Methods**

#### **Participants and Clinical Outcome**

SCZ-NA cohort comprised individuals with predominantly recent-onset SCZ. For samples harmonized from the NAPLS study, we included only clinical high-risk youth from NAPLS 2 or NAPLS 3 who subsequently developed a schizophrenia spectrum disorder (N=139)^[1]^. In SCZ-NA, SCZ diagnosis and AOO was determined using the psychosis module of the Structured Clinical Interview for DSM Disorders or the Kiddie Schedule for Affective Disorders and Schizophrenia. For a subset of participants, AOO was calculated based on date of birth and the date at which participants reported experiencing full-blown psychotic symptoms for more than one week ^[2]^. In SCZ-UKBB, SCZ diagnoses were determined through self-reported diagnoses, ICD-10 diagnoses from hospital admission records, death records listing schizophrenia as a cause of death, and primary care records. AOO was defined from two sources to maximize sample size, taking the earlier value of: (1) self-reported age at diagnosis and (2) age at first unusual/psychotic symptoms.

#### **Genotyping and imputation**

For SCZ-NA. DNA was extracted from whole blood, saliva, or buccal samples and genotyped in two batches on the Illumina Global Screening Array (700,078 markers). Genotypes were processed using RICOPILI ^[3]^, including pre-imputation variant and sample-level QC. Imputation was performed using the Trans-Omics for Precision Medicine (TOPMed) reference panel ^[4–6]^. Post-imputation variant-level QC retained SNPs with minor allele frequency (MAF) > 0.01 and imputation INFO > 0.8; variants in the extended MHC region were excluded with the exception of the single SNP in that region showing the strongest association with SCZ ^[7,8]^. The final dataset comprised 5,991,083 high-quality SNPs for PRS calculation. For SCZ-UKBB, we restricted analyses to participants whose genetic data passed UK Biobank genotyping QC^[9]^. Analyses proceeded from the imputed genotype dataset. Applying the same exclusion rules yielded 7,103,933 SNPs for PRS analyses.

#### **Pathway Definition and SNP-to-gene mapping**

Pathway-specific SCZ-PRSs were computed using the 18 mutually exclusive neurodevelopmental gene-sets previously derived from BrainSpan transcriptomic data using weighted gene co-expression network analysis (WGCNA)^[10]^. To assign SNPs to genes within each pathway, we applied 13 distinct SNP-to-gene mapping definitions. Mapping definitions differed with respect to both: (1) how variants located within gene boundaries were handled; and (2) how non-coding variants outside gene bodies were linked to genes.

***Variants Located within Gene Boundaries.*** For SNPs located between the transcription start and end sites of genes, we considered two genic annotation filters:

1. Exon + Splice: variants annotated by ANNOVAR as “exonic,” “splicing,” “ncRNA_exonic;splicing,” “exonic;splicing,” “ncRNA_exonic,” or “ncRNA_splicing.”
2. Exon + Intron: all variants located between the transcription start and end sites of a gene, including both exonic and intronic variants (i.e., and therefore inherently including splicing site variants).

##### ***Non-Coding Variants.*** For non-coding SNPs outside gene bodies, one class of mappings relied on physical proximity, or distance-based mappings to gene boundaries; the other used functional annotations.

**Distance-based mapping definitions.** For physical proximity non-coding SNP-to-gene mappings, we used a common definition of 35-kb upstream and 10-kb downstream of the transcription start and end site, respectively, of each gene. Furthermore, we implemented three versions of this distance-based strategy that differed in how they handled variants within the target gene’s distance window that overlapped neighboring genes:

1. “Exon_Splice_35kb_10kb_non_coding”: included target-gene exonic/splice variants and non-coding variants within the 35-kb upstream/10-kb downstream window, but excluded SNPs if they were located in another gene’s exons or splice sites.
2. “Exon_Intron_35kb_10kb_non_coding”: included target-gene exonic/intronic variants and non-coding variants (i.e., annotated by Annovar as “intergenic,” “upstream,” “downstream,” or “upstream;downstream”) within the 35-kb upstream/10-kb downstream window, but excluded SNPs if they were located in another gene’s exons or splice sites.
3. “Exon_Intron_35kb_10kb_All”: included target-gene exonic/intronic variants and all coding and non-coding variants within the 35-kb upstream/10-kb downstream window, even if the SNP overlapped another gene’s exonic, intronic, or splice-site regions.

**Functional annotation-based mapping definitions.** A second class of mapping definitions linked non-coding variants to genes using functional genomic evidence. We evaluated four types of functional mapping sources:

1. Brain-specific expression and splicing quantitative trait loci (eQTL/sQTLs). eQTLs link genetic variants to overall gene expression levels, whereas sQTLs link variants to alternative splicing patterns that can generate different isoforms and protein forms. We used all brain QTLs from GTEx ^[11]^ ,which leveraged whole-genome sequence data from 838 postmortem donors and included 17,382 RNA-seq samples from 948 donors overall, and PsychENCODE ^[12]^, which identified QTLs in 1,387 genotype-expression-matched adult prefrontal cortex samples;
2. Cross-tissue gene enhancers and promoters from GeneHancer^[13]^(GH) which non-redundantly merged >1M candidate regulatory elements into ~250k scored enhancers and assigned enhancer-gene links using integrated evidence; for GH-based SNP-to-gene mapping, we included only enhancer-gene links supported by both an elite enhancer (an enhancer supported by at least two evidence sources) and an elite gene–enhancer association (an association supported by at least two methods).
3. Fetal brain 3D chromatin contacts derived from paracentral cortex of three individuals of gestation week 17-18 ^[14]^;
4. Union of all four functional annotation sources.

For each of these four functional sources, we generated two pathway definitions by pairing the functional annotations with the two genic filters described above (Exon + Splice and Exon + Intron), yielding 10 functional annotation-based SNP-to-gene mapping definitions. Together with the 3 distance-based mappings, this produced 13 total SNP-to-gene definitions per pathway and 234 pathway-specific SCZ-PRSs per cohort (18 pathways × 13 mappings).

#### **Polygenic risk scores calculation and ancestry adjustment**

To account for continuous variation in genetic ancestry, we adopted a state-of-the-art dimensional approach that adjusts PRS across continuous axes of genetic ancestry, by accounting for differences in both the mean and variance of the PRS distribution across populations ^[15]^. For each cohort, we projected individuals into the 1000 Genomes reference PCA space ^[16]^ and constructed SCZ-PRS for 1000 Genomes subjects using overlapping SNPs, using Bayesian shrinkage to estimate posterior SNP weights. As implemented in a previously published pipeline^[15]^, we modeled ancestry-related mean and variance effects by regressing these 1000 Genomes SCZ-PRSs on the first five PCs and regressing squared residuals on the same PCs, then converted cohort PRS to ancestry-adjusted Z-scores using the PC-predicted mean and spread. This approach adjusts for ancestry-related shifts in central tendency and dispersion without excluding admixed individuals ^[15]^. In the SCZ-NA cohort, it improved case-control prediction by >1% proportion of variance explained relative to categorical-based within-ancestry Z-scoring. In the SCZ-UKBB cohort, which is of predominantly European ancestry, it still produced a 0.3% increase in variance explained. Together, these findings support this ancestry adjustment as the preferred option for maximizing PRS accuracy and statistical power in samples spanning diverse ancestries.

#### **Statistical Analysis**

For validation analyses relating genome-wide SCZ-PRS to SCZ case-control status, R^2^ was estimated using the *jointScoreTest()* method implemented in the GENESIS package^[17]^. PRS optimization and performance testing were conducted in the same sample. Accordingly, the reported R^2^ and p-values should be interpreted as within-sample estimates and not as unbiased measures of out-of-sample predictive performance in independent cohorts.
